# Perceived quality of care among households ever enrolled in a community-based health insurance scheme in two districts of northeast Ethiopia: a multilevel analysis

**DOI:** 10.1101/2021.10.18.21265144

**Authors:** Mohammed Hussien, Muluken Azage, Negalign Berhanu Bayou

## Abstract

**Objectives:** The purpose of this study was to examine how clients perceived about the quality of health care they received and to identify associated factors at the individual and facility-level.

**Design:** A community-based, cross-sectional study

**Setting:** Health centers in two districts

**Participants:** 1081 rural households who had ever been enrolled in a community-based health insurance and had visited a health center at least once in the previous 12 months, as well as 194 health care providers working in 12 health centers.

**Outcome measures:** The outcome variable of interest was the perceived quality of care, which was measured using a 17-item scale. Respondents were asked to rate the degree to which they agreed on 5-point response items relating to their experiences with health care in the outpatient departments of nearby health centers. A multilevel linear regression analysis was used to identify predictors of perceived quality of care.

**Results:** The mean perceived quality of care was 70.28 (SD=8.39). Five dimensions of perceived quality of care were extracted from the factor analysis, with the patient-provider communication dimension having the highest mean score (M=77.84, SD=10.12), and information provision having the lowest (M=64.67, SD=13.87). Wealth status, current insurance status, perceived health status, presence of chronic illness, time to a recent health center visit, work experience of health care providers and patient volume were the factors significantly associated with perceived quality of care. An interaction term between patient volume and staff job satisfaction also showed significant association.

**Conclusions:** Much work remains to improve the quality of care, especially on information provision and access to care quality dimensions. A range of individual and cluster-level characteristics influence the perceived quality of care. For a better quality of care, it is vital to optimize the patient-provider ratio, and enhance staff job satisfaction.

**Strengths and limitations of this study:**

▸ The study tried to assess the quality of care from the clients’ point of view using a validated multidimensional scale.
▸ This is the first cross-sectional study in Ethiopia, which considered health center (cluster) level variables that have association with perceived quality of care.
▸ We tested for the existence of endogeneity between current insurance status and quality of care. Although the results indicated no evidence of endogeneity, it is still possible due to omitted variables. Active insurance members may report a higher perception score quality of care as a result of their desire to stay in the scheme.
▸ Because of the cross-sectional nature of the study, it is impossible to establish a cause-and-effect relationship.

## INTRODUCTION

Health care providers and patients define quality of care differently and attach varying levels of importance to its attributes. When assessing the quality of care, health care professionals tend to prioritize technical competence, while patients place a high value on patient-centeredness, amenities, and reputation.^1^ The emphasis on health care quality measurement has shifted away from the viewpoints of health care providers to people-centered approaches that rely on patient perceptions.^2-4^ Patients’ perception of health care quality has become an essential element of quality measurement due to its link with health service utilization. It is based on a mix of patient experiences, processed information and rumors.^5^ Patient experience surveys elicit data on the transactional components of care, which are process-related, as well as the interpersonal interactions that occur over the course of care.^6^ Individuals receiving care are asked about their experiences of health facility encounters to report if particular processes or events occurred.^7^ Patient experience measurements have received increased attention and are widely employed to inform quality improvement, and pay-for-performance.^8^ Patient experience is consistently and positively associated with patient safety and clinical effectiveness, adherence to prevention and treatment recommendations, and technical quality of care.^9 10^

Quality of health care is vital to the success of universal health coverage (UHC) initiatives, like community-based health insurance (CBHI). The development of CBHI schemes must be accompanied by improvements in the quality of care.^11 12^ To build sustainable CBHI schemes, members must believe that the benefits of health care provided via health insurance coverage outweigh the benefits of not being insured.^13^ Patients’ positive experiences with the quality of care provided under insurance schemes increase their trust in the health system and insurance schemes.^14 15^ As a result, they are more likely to use health care services and participate in health insurance plans.^16^ If insured clients are unable to access high quality services, they lose trust in service providers and seek care elsewhere,^17^ making them less likely to pay premiums.^18 19^

The ultimate goal of UHC is to ensure that all people who need health services receive high quality care without financial strain.^20^ Although increased health care coverage is promising with the implementation of CBHI, quality of care remains a key impediment to achieving UHC.^20 21^ Increasing access to essential health services without improving their quality would not bring the intended health outcomes.^2 4^ For example, more than eight million deaths amenable to a high quality of care occurred in low- and middle-income countries, making poor-quality of care a bigger obstacle to mortality reduction than lack of access to care.^21^

Poor quality of care is also a major issue that jeopardizes the long-term viability of many CBHI schemes.^11 22^ Findings of systematic reviews revealed that the quality of care was a key factor that influenced enrollment and renewal decisions of CBHI membership.^23 24^ Some quality concerns include ‘unavailability and perceived poor quality of prescribed medicines, misbehavior of health professionals, and the differential treatment of the insured in favor of the uninsured patients, unclean hospital environment, long queues, lack of diagnostic equipment, and long waiting hours to obtain health care’.^24^

To promote optimal utilization, stable finance, and better outcomes, the quality of health care must be monitored on a regular basis.^17^ Previous studies in Ethiopia focused on surveys of client satisfaction and did not employ multidimensional measurement scales.^25 26^ To our knowledge, the quality of care delivered under the CBHI in Ethiopia has never been investigated using multidimensional metrics from the perspective of service users at the community level. There is also a paucity of literature on facility-level variables that influence the quality of care. Therefore, the purpose of this study was to examine the perceived quality of care (PQoC) from the perspective of clients, and identify associated factors at the individual and facility-level.

Improving quality of care and CBHI are among Ethiopia’s top priorities in its health sector strategic plan.^27^ The findings of this study will inform relevant stakeholders on the current state of clients’ perceptions of the quality of care, and will be an essential input for quality improvement initiatives. It will also provide useful information for decision-makers to address challenges in the country’s endeavors to establish higher-level insurance pools.

## METHODS

### Study setting and population

A community-based cross-sectional study was conducted in rural parts of two neighboring districts in northeast Ethiopia, Tehulederie and Kallu. Tehulederie is divided into 20 rural and seven urban Kebeles (subdistricts) with a population of 145,625, of which 87.5% reside in rural areas. There are five health centers and one primary hospital in the district. It was one of the 13 districts in Ethiopia where CBHI was piloted in 2011. The scheme was introduced in Kallu district after two years, in July 2013. Kallu is divided into 36 rural and four urban Kebeles, and has nine health centers. It is the most populous district in the zone, with a population of 234,624, of which 89.11% live in the rural area.^28^

The study population of interest were rural households who had ever been enrolled in the CBHI scheme before January 2020. To minimize recall bias, households who had not used health care in the 12-month period before data collection were excluded from the study. The sample size was calculated using MedCalc software by assuming a mean difference of two independent groups. A previous study on PQoC reported mean scores of 5.2 and 5.4 with standard deviations (SD) of 0.8 and 0.7 among insured and uninsured respondents, respectively.^29^ Using this output and assuming an 80% power, 95% confidence level and equally sized groups, a sample size of 446 was calculated. Considering a design effect of 1.5 attributable to multi-stage sampling and a potential non-response rate of 10%, the effective sample size was estimated to be 736 households. Alternative sample size of 1257 was calculated for a companion article as part of a research project examining the sustainability of a CBHI in Ethiopia. Among those, 1081 eligible households participated in this study. Furthermore, 194 health care providers from 12 health centers participated in the study to provide cluster-level data.

### Data collection and measurement

The data were collected from 04 February to 21 March 2021. The study participants were recruited using a three-level multistage sampling approach. First, 12 clusters of Kebeles organized under a health center catchment area were selected. Then, 14 rural Kebeles were drawn randomly using a lottery method proportional to the number of Kebeles under each cluster. Accordingly, five Kebeles from Tehulederie and nine from Kallu were included. A list of households who have ever been enrolled in the CBHI was obtained from the membership registration logbook of each Kebele. The required sample was generated at random from each Kebele, proportional to the number of households who have ever been enrolled in the scheme, using random number generator software.

Individual-level data was collected through face-to-face interviews with household heads at their homes or workplace using a structured questionnaire via an electronic data collection platform. The data collectors submit the completed forms to a data aggregating server on a daily basis, which allowed us to review the submissions and streamline the supervision process.

The PQoC, which is the outcome variable of interest, was measured using a 17-item scale designed after a thorough review of validated tools.^29-33^ Respondents were asked to rate the extent to which they agreed on a set of items relating to their experiences with the health care they received in the outpatient departments of nearby health centers. Each item was designed on a 5-point response format, with 1=strongly disagree, 2=disagree, 3=neutral, 4=agree and 5=strongly agree. The summary scores for the PQoC and its dimensions were calculated for individual respondents by adding the scores for each item. This gives a scale ranging from 17 (1×17) to 85 (5×17) for the overall PQoC score. When reporting the results, the scores were arithmetically transformed to a scale of 20 to 100.^34^ This allows the comparison of mean scores of PQoC with its dimensions, and measurement items on a common scale.

Wealth index was generated using the principal component analysis method. The scores for 15 types of assets were translated into latent factors, and a wealth index was created based on the first factor that explained most of the variation. The study households were grouped into wealth tertile – lower, medium and higher based on the index. Perceived health status was measured based on a household head’s subjective assessment of the health status of the household, and was rated as “poor, fair, good, very-good, or excellent”. However, for analysis purpose, it was recategorized into “fair, good, and very-good”, by merging the two extreme response categories to the next option due to fewer replies.

Before the data collection, the questionnaire was pre-tested on a sample of 84 randomly selected participants in one Kebele. As part of the pre-test, a cognitive interview was conducted on selected items using the verbal probe technique among eight respondents to determine if the items and response categories were understood, and interpreted by the potential respondents as intended. Accordingly, the phrasing of some items and response options were modified, and some items were omitted.

Cluster-level data were collected from 12 health centers that provide health care for the population in the sampled Kebeles. Patient volume data were obtained by reviewing the monthly service delivery reports of health centers, while data related to work experience, affective commitment and job satisfaction were collected through a self-administered questionnaire among health care providers who worked more than one year in the current facility.

Patient volume was measured using the daily average number of patients managed by a health care provider in the outpatient department. It was calculated by dividing the number of patients who visited the health center in the last six months before the study by the number of working days, and then by the number of consultation rooms in each health center.^35^ Affective commitment and job satisfaction were composite variables which were assessed using a 5-point Likert scale. Affective commitment was measured with a seven-item questionnaire based on a modified version of the Meyer et al. scale, which had previously been used in a hospital setup.^36^ Staff job satisfaction was measured using a 10-item scale, which was adapted from a previous study among health care workers in Ethiopia.^37^ Average affective commitment and job satisfaction scores were computed for each health center.

### Data analysis

The data were analyzed using Stata version 17.0. Exploratory factor analysis was performed to assess the validity of the quality measurement scale. The Bartlett’s test of Sphericity and Kaiser-Mayer-Olkin’s (KMO) measure of sampling adequacy were performed to assess the appropriateness of the data for factor analysis. The principal component factor method of extraction and Promax rotation with Kaiser Normalization was used. The Eigenvalue greater than one decision rule was used to determine the appropriate number of factors to be extracted. Items with loadings below 0.40 were removed from the analysis.^38^ Correlation coefficients were used to test construct validity. Item-total score correlation, dimension-total score correlation and dimension intercorrelation were computed. The total score was the mean score of the ratings for all items of the scale, and the dimension score was the factor scores. A questionnaire has good construct validity when the item-total score correlations are higher than 0.40, dimension intercorrelations are less than 0.80, and dimension-total score correlations are higher than dimension intercorrelations.^31^ Cronbach’s alpha coefficients were generated for each dimension to assess the internal consistency. Reliability of the scale was considered acceptable if Cronbach’s alpha coefficient was 0.60 or higher.^38^

To compare mean scores of PQoC and its dimensions among subgroups, an independent t-test and a one-way analysis of variance (ANOVA) with Tukey’s post-hoc test were used. Because the outcome variable was considered as a continuous variable, a multilevel linear regression model was fitted to identify its predictors. The Restricted Maximum Likelihood estimation approach was used because it is appropriate for smaller cluster sizes.^39^ The PQoC was assumed to be influenced by the characteristics of households (individual-level variables) as well as the characteristics of health centers (cluster-level variables). Cluster-level data were linked to individual-level data based on the usual source of health care for each study participant. Considering the hierarchical structure of the data, where patients are nested within health centers, a two-level linear regression model was applied. Four models were estimated to choose the one that best fits the data. The first model, or the null model (a model without predictors) is given by:

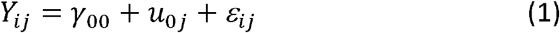

The null model estimates three parameters: the average intercept (*Y*_*00*_), the between health center error, or deviation, from the average intercept (*u*_*0j*_), and the individual-level residual, or variation in individual scores within health centers (*ε*_*ij*_). The second model estimated PQoC (*Y*_*ij*_) for individual household *i* at health center *j*. We treat PQoC as a function of a matrix of individual-level variables (*X*_*ij*_) which include age, gender, education and marital status of the household head; wealth status; household size; current health insurance status; presence of chronic illness in the household; perceived health status, and time to a recent visit to a health center, and expressed as:

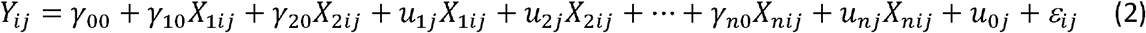

where *u*_*1j*,_ *u*_*2j…*_*u*_*nj*_ indicate the random error terms connected to each *X*_*ij*_.

The third model estimated the PQoC as a function of cluster-level variables (Z_*j*_) that include average work experience, affective commitment and job satisfaction of health care providers, and patient volume. The model takes into account the differences between health centers and explains these differences in terms of these characteristics. It is given by:

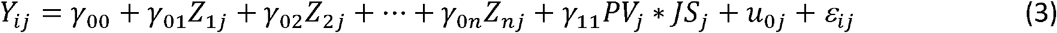

where *PV*_*j*_*\*JS*_*j*_ indicates an interaction term between patient volume and job satisfaction in which job satisfaction was assumed to moderate the effect between patient volume and PQoC. The interaction effect was tested by plotting the marginal effects of interaction terms. The two variables were centered towards the grand mean to facilitate the interpretation of the coefficients.

By combining model II and III, the fourth model estimated the PQoC as a function of both individual and cluster-level variables, and can be written as:

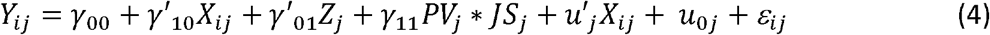

where γ_10_ and γ_01_ are the vector of coefficients of n explanatory variables whose values are at *X*_*1ij*_, *X*_*2ij*_, …, *X*_*nij*_ for the *i*^*th*^ individual within the *j*^*th*^ cluster, and *Z*_*1j*_, *Z*_*2j*_,, *Z*_*nj*_ for the *j*^*th*^ cluster, respectively. The intercept γ_00_ and slopes *γ*_*01*_, *γ*_*10*_ and *γ*_*11*_ are fixed effects, while *u*_*0j*_, *u*_*j*_ and *ε*_*ij*_ are random effects.

This multilevel regression decomposes the total variances into two independent components: 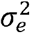, which is the variance of individual-level errors *ε*_*ij*_, and 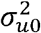, which is the variance of cluster-level errors *u*_*oj*_. From this model we can define the intraclass correlation (ICC) by the equation:^40^

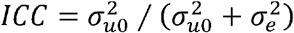

The ICC and proportional change in variance (PCV) were used to report the measures of variation (random effects). The need for multilevel analysis, which considers cluster-level factors, was tested using the ICC. The ICC shows the variation in PQoC accounted for cluster-level characteristics. Statistically significant variability between health centers justifies the need to consider cluster-level factors.^41^ The PCV expresses the change in the cluster-level variance between the empty model and models with more terms, and is calculated by PCV = (V_A_ - V_B_)/V_A_, where V_A_ = variance of the null model, and V_B_ = variance of the model with more terms. It measures the total variation explained by individual and cluster-level factors.

The measures of association (fixed-effects) estimate the association between the PQoC score and various explanatory variables. The existence of a statistically significant association was determined at p-values of <0.05. The degree of the association was assessed using regression coefficients, and their statistical significance was determined at a 95% confidence interval. Models were compared using the Deviance Information Criteria (DIC) and Akaike Information Criteria (AIC). The best fit model was determined to have the lowest DIC and AIC values. The preliminary analysis confirmed no violation of the assumptions of normality, linearity, homoscedasticity, and multicollinearity. The presence of multicollinearity was determined using Variance Inflation Factor with a cutoff point of 5.

### Patient and Public Involvement

No patient involved

## RESULTS

### Background characteristics of the study population

The household survey included 1081 respondents who had visited a health center at least once in the previous 12 months prior to the study. The average age of the study participants was 49.25 years (SD=12.07), with slightly more than half (51.34%) were between the age ranges of 45 and 64, and 12.67% being 65 and older. Of the total household heads, 938 (86.77%) were men, and 1003 (92.78%) were currently married. One-fifth of the study participants (20.91%) attended formal education, and 62.72% had a household size of five or above.

Nearly ninety percent of the households (87.14%) were active members of the CBHI scheme at the time of the study. A quarter of households (25.72%) had one or more individuals with a known chronic illness informed by a healthcare provider. One-third of respondents (33.58%) rated their household health status as very-good, while 207 (19.15%) and 511 (47.27%) rated it as fair and good, respectively. Nearly half of the households (46.16%) had visited a health center within three months prior to the study, while 31.73% and 22.11% had their most recent visit to a health center before 6-12 and 3-6 months, respectively (Table 1).

**Table 1:** Independent t-test and one-way ANOVA comparing mean scores of the PQoC (20-100 scale) across respondent characteristics in two districts of northeast Ethiopia, 2021

| Variable | Categories | N=1081 | % | PQoC score |  | t/F-test |
| --- | --- | --- | --- | --- | --- | --- |
|  |  |  |  | M | SD |  |
| Age in years | 25 – 44 | 389 | 35.99 | 69.97 | 7.78 | 1.08 |
|  | 45 – 64 | 555 | 51.34 | 70.26 | 8.52 |  |
|  | 65+ | 137 | 12.67 | 71.20 | 9.49 |  |
| Gender | Men | 938 | 86.77 | 70.15 | 8.21 | -1.31 |
|  | Women | 143 | 13.23 | 71.13 | 9.51 |  |
| Marital status | Divorced/widowed | 78 | 7.22 | 71.61 | 10.95 | 1.46 |
|  | Married | 1003 | 92.78 | 70.17 | 8.16 |  |
| Attend formal education | No | 855 | 79.09 | 70.29 | 8.48 | 0.07 |
|  | Yes | 226 | 20.91 | 70.24 | 8.05 |  |
| Household size | < Five | 403 | 37.28 | 70.85 | 8.63 | 1.73 |
|  | ≥ Five | 678 | 62.72 | 69.94 | 8.25 |  |
| Wealth tertile | Lowest | 361 | 33.40 | 71.77 | 9.15 | 8.83<sup>#</sup> |
|  | Medium | 360 | 33.30 | 69.36<sup>b</sup> | 8.16 |  |
|  | Highest | 360 | 33.30 | 69.70<sup>b</sup> | 7.62 |  |
| Current insurance status | Ex-member | 139 | 12.86 | 67.66 | 9.65 | -3.96<sup>#</sup> |
|  | Active-member | 942 | 87.14 | 70.66 | 8.13 |  |
| Perceived health status | Fair | 207 | 19.15 | 72.28 | 8.84 | 8.04<sup>#</sup> |
|  | Good | 511 | 47.27 | 70.08<sup>b</sup> | 7.83 |  |
|  | Very-good | 363 | 33.58 | 69.41<sup>b</sup> | 8.73 |  |
| Chronic illness | No | 803 | 74.28 | 69.54 | 8.29 | -4.96<sup>#</sup> |
|  | Yes | 278 | 25.72 | 72.40 | 8.33 |  |
| Last health center visit | < 3 months | 499 | 46.16 | 70.75<sup>b</sup> | 8.99 | 4.78<sup>§</sup> |
|  | 3-6 months | 239 | 22.11 | 70.94<sup>b</sup> | 7.60 |  |
|  | 6-12 months | 343 | 31.73 | 69.13 | 7.92 |  |
| Total |  | 1081 | 100 | 70.28 | 70.28 |  |

Statistical significance for t-test/F-test is indicated by <sup>§</sup> $p < 0.01$ , and <sup>#</sup> $p < 0.001$ . Based on Tukey's post-hoc test, mean values sharing letter 'b' are not significantly different in the group at the 5% level.

The median work experience of health care providers involved in this study ranges from three to ten years. The mean scores of affective commitment and job satisfaction were 29.00 and 30.95 (SD=2.08 and 3.17), respectively. The average patient volume was 32.17 per day per care provider, with a range of 19 to 43 (SD=7.83).

### Factor analysis

Sampling was adequate as measured by the KMO (0.83), and Bartlett’s test of sphericity was significant (p<0.001). Two items were removed from further analysis due to loadings below 0.40, and one item was removed due to low communality. The factor analysis extracted five dimensions that explained 59.25% of the total variation (online supplemental file 1). The item-total score correlations ranged from 0.268 to 0.622, four items had correlations less than 0.40. The dimension intercorrelations varied from 0.031 to 0.434, all of which were less than the 0.80 criterion, indicating that each dimension was distinct enough to be considered an independent measure. Dimension-total score correlation ranged between 0.417 to 0.772, all significant at a p-value of 0.001, and were higher than dimension intercorrelations. The scale was tested for reliability and had an overall Cronbach’s alpha coefficient of 0.804. The Cronbach’s alpha coefficients for the five dimensions exceeded 0.60, except for the access to care subscale, which had an alpha coefficient of 0.531.

### Perceived quality of care

The minimum and maximum PQoC scores were 37.65 and 97.65, respectively. The mean score was 70.28 (95% CI: 69.77, 70.78) with an SD of 8.39. The aggregated mean score at the health center-level ranges from 64.94 to 74.06. Patient-provider communication had the highest mean score (M=77.84, SD=10.12) of the five quality dimensions, while information provision had the lowest score (M=64.67, SD=13.87). The mean score for each measurement item is summarized by online supplemental file 2.

An independent t-test and a one-way ANOVA were performed to compare the mean scores of PQoC and its dimensions between subgroups. As shown under Table 1, there was a significant difference in the PQoC mean score for wealth tertile at p<0.05 (F=8.83, p=0.001). Tukey’s post-hoc test indicated that the mean score of PQoC for the lowest wealth tertile (M=71.77, SD=9.15) was significantly different from both the medium (M=69.36, SD=8.16) and highest (M=69.70, SD=7.62) wealth tertile. However, no significant difference was seen between medium and high wealth tertile. The ANOVA test also showed that the PQoC mean score showed significant differences based on the respondents perceived health status and time to a recent visit to a health center, with (F=8.04, p<0.001) and (F=4.78, p<0.01), respectively. There was a significant difference in the mean score of PQoC between active insurance members (M=3.53, SD=0.41) and ex-members (M=3.38, SD=0.48); t = 3.96, p<0.001. The mean PQoC score of households with chronic illness (M=3.62, SD=0.42) was also significantly higher compared to those who did not have chronic illness (M=3.48, SD=0.42); t = 4.95, p<0.001. The results of an independent t-test and a one-way ANOVA that compare the differences in mean scores of the five dimensions between subgroups is displayed by Table 2.

**Table 2:**
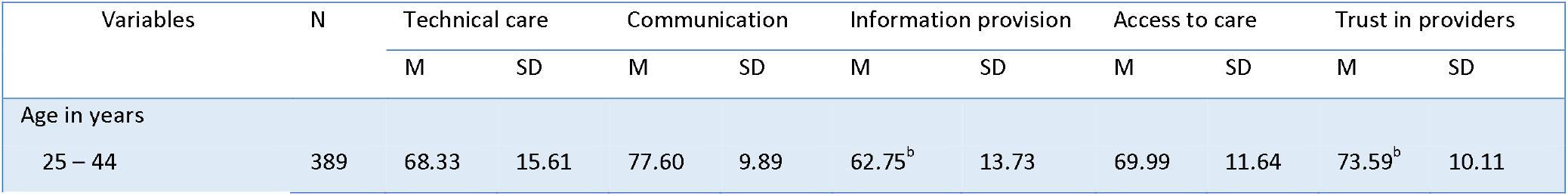
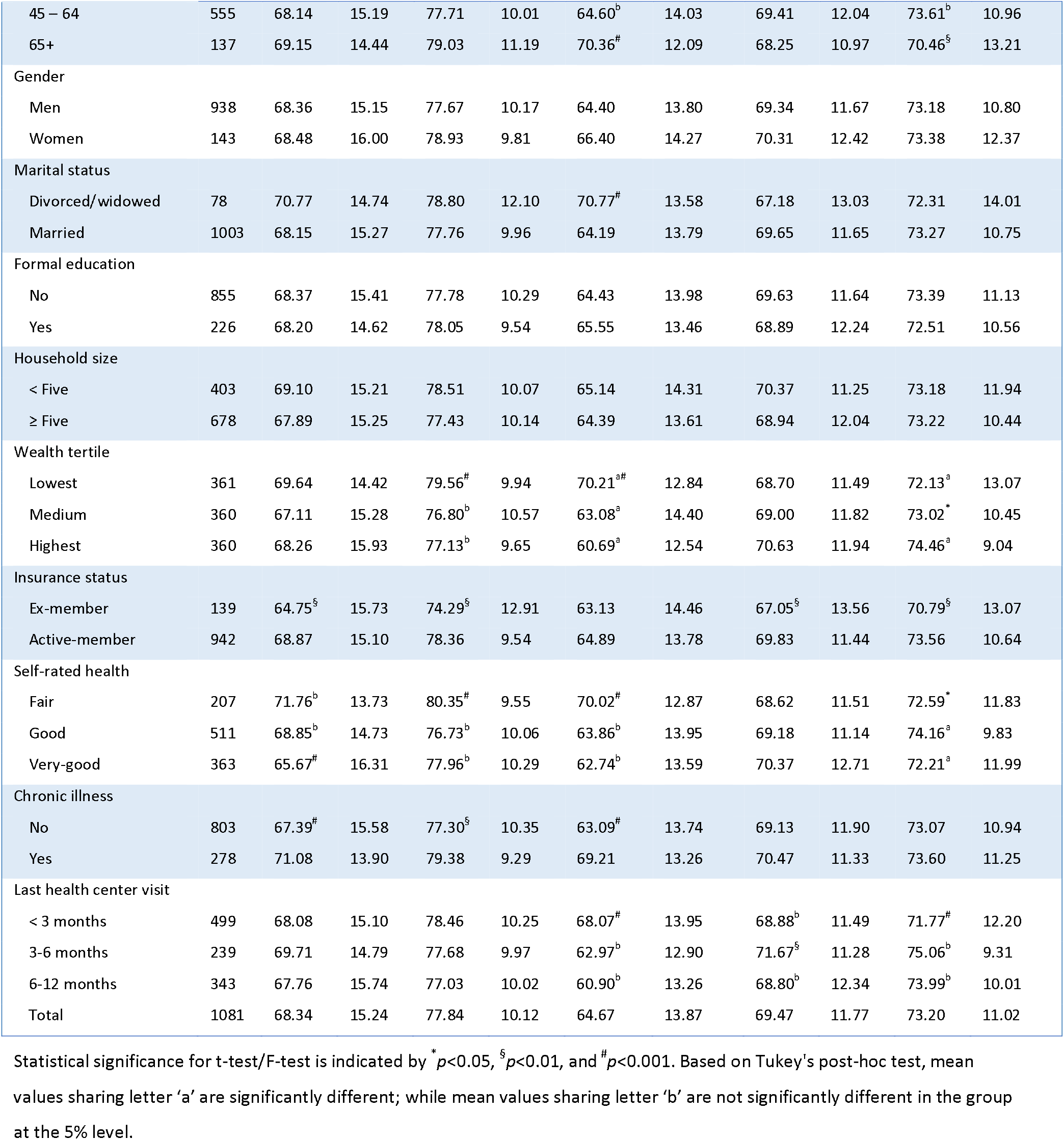
Independent t-test and one-way ANOVA comparing mean scores of PQoC dimensions (20-100 scale) across respondent characteristics in two districts of northeast Ethiopia, 2021

| Variables | N | Technical care |  | Communication |  | Information provision |  | Access to care |  | Trust in providers |  |
| --- | --- | --- | --- | --- | --- | --- | --- | --- | --- | --- | --- |
|  |  | M | SD | M | SD | M | SD | M | SD | M | SD |
| Age in years |  |  |  |  |  |  |  |  |  |  |  |
| 25 – 44 | 389 | 68.33 | 15.61 | 77.60 | 9.89 | 62.75<sup>b</sup> | 13.73 | 69.99 | 11.64 | 73.59<sup>b</sup> | 10.11 |
| 45 – 64 | 555 | 68.14 | 15.19 | 77.71 | 10.01 | 64.60<sup>b</sup> | 14.03 | 69.41 | 12.04 | 73.61<sup>b</sup> | 10.96 |
| 65+ | 137 | 69.15 | 14.44 | 79.03 | 11.19 | 70.36<sup>#</sup> | 12.09 | 68.25 | 10.97 | 70.46<sup>§</sup> | 13.21 |
| Gender |  |  |  |  |  |  |  |  |  |  |  |
| Men | 938 | 68.36 | 15.15 | 77.67 | 10.17 | 64.40 | 13.80 | 69.34 | 11.67 | 73.18 | 10.80 |
| Women | 143 | 68.48 | 16.00 | 78.93 | 9.81 | 66.40 | 14.27 | 70.31 | 12.42 | 73.38 | 12.37 |
| Marital status |  |  |  |  |  |  |  |  |  |  |  |
| Divorced/widowed | 78 | 70.77 | 14.74 | 78.80 | 12.10 | 70.77<sup>#</sup> | 13.58 | 67.18 | 13.03 | 72.31 | 14.01 |
| Married | 1003 | 68.15 | 15.27 | 77.76 | 9.96 | 64.19 | 13.79 | 69.65 | 11.65 | 73.27 | 10.75 |
| Formal education |  |  |  |  |  |  |  |  |  |  |  |
| No | 855 | 68.37 | 15.41 | 77.78 | 10.29 | 64.43 | 13.98 | 69.63 | 11.64 | 73.39 | 11.13 |
| Yes | 226 | 68.20 | 14.62 | 78.05 | 9.54 | 65.55 | 13.46 | 68.89 | 12.24 | 72.51 | 10.56 |
| Household size |  |  |  |  |  |  |  |  |  |  |  |
| < Five | 403 | 69.10 | 15.21 | 78.51 | 10.07 | 65.14 | 14.31 | 70.37 | 11.25 | 73.18 | 11.94 |
| ≥ Five | 678 | 67.89 | 15.25 | 77.43 | 10.14 | 64.39 | 13.61 | 68.94 | 12.04 | 73.22 | 10.44 |
| Wealth tertile |  |  |  |  |  |  |  |  |  |  |  |
| Lowest | 361 | 69.64 | 14.42 | 79.56<sup>#</sup> | 9.94 | 70.21<sup>a#</sup> | 12.84 | 68.70 | 11.49 | 72.13<sup>a</sup> | 13.07 |
| Medium | 360 | 67.11 | 15.28 | 76.80<sup>b</sup> | 10.57 | 63.08<sup>a</sup> | 14.40 | 69.00 | 11.82 | 73.02<sup>*</sup> | 10.45 |
| Highest | 360 | 68.26 | 15.93 | 77.13<sup>b</sup> | 9.65 | 60.69<sup>a</sup> | 12.54 | 70.63 | 11.94 | 74.46<sup>a</sup> | 9.04 |
| Insurance status |  |  |  |  |  |  |  |  |  |  |  |
| Ex-member | 139 | 64.75<sup>§</sup> | 15.73 | 74.29<sup>§</sup> | 12.91 | 63.13 | 14.46 | 67.05<sup>§</sup> | 13.56 | 70.79<sup>§</sup> | 13.07 |
| Active-member | 942 | 68.87 | 15.10 | 78.36 | 9.54 | 64.89 | 13.78 | 69.83 | 11.44 | 73.56 | 10.64 |
| Self-rated health |  |  |  |  |  |  |  |  |  |  |  |
| Fair | 207 | 71.76<sup>b</sup> | 13.73 | 80.35<sup>#</sup> | 9.55 | 70.02<sup>#</sup> | 12.87 | 68.62 | 11.51 | 72.59<sup>*</sup> | 11.83 |
| Good | 511 | 68.85<sup>b</sup> | 14.73 | 76.73<sup>b</sup> | 10.06 | 63.86<sup>b</sup> | 13.95 | 69.18 | 11.14 | 74.16<sup>a</sup> | 9.83 |
| Very-good | 363 | 65.67<sup>#</sup> | 16.31 | 77.96<sup>b</sup> | 10.29 | 62.74<sup>b</sup> | 13.59 | 70.37 | 12.71 | 72.21<sup>a</sup> | 11.99 |
| Chronic illness |  |  |  |  |  |  |  |  |  |  |  |
| No | 803 | 67.39<sup>#</sup> | 15.58 | 77.30<sup>§</sup> | 10.35 | 63.09<sup>#</sup> | 13.74 | 69.13 | 11.90 | 73.07 | 10.94 |
| Yes | 278 | 71.08 | 13.90 | 79.38 | 9.29 | 69.21 | 13.26 | 70.47 | 11.33 | 73.60 | 11.25 |
| Last health center visit |  |  |  |  |  |  |  |  |  |  |  |
| < 3 months | 499 | 68.08 | 15.10 | 78.46 | 10.25 | 68.07<sup>#</sup> | 13.95 | 68.88<sup>b</sup> | 11.49 | 71.77<sup>#</sup> | 12.20 |
| 3-6 months | 239 | 69.71 | 14.79 | 77.68 | 9.97 | 62.97<sup>b</sup> | 12.90 | 71.67<sup>§</sup> | 11.28 | 75.06<sup>b</sup> | 9.31 |
| 6-12 months | 343 | 67.76 | 15.74 | 77.03 | 10.02 | 60.90<sup>b</sup> | 13.26 | 68.80<sup>b</sup> | 12.34 | 73.99<sup>b</sup> | 10.01 |
| Total | 1081 | 68.34 | 15.24 | 77.84 | 10.12 | 64.67 | 13.87 | 69.47 | 11.77 | 73.20 | 11.02 |

Statistical significance for t-test/F-test is indicated by \* $p < 0.05$ , § $p < 0.01$ , and # $p < 0.001$ . Based on Tukey's post-hoc test, mean values sharing letter 'a' are significantly different; while mean values sharing letter 'b' are not significantly different in the group at the 5% level.

The mean PQoC score was significantly different among health centers (F=11.85, p<0.001). The mean scores for the five dimensions were also significantly different among health centers at p<0.001 level: technical care (F=8.66), patient-provider communication (F=6.65), information provision (F=47.42), access to care (F=36.87) and trust in care providers (F=6.98). The mean scores of the PQoC and its dimensions across the 12 health centers are depicted using a radar chart (Figure 1). The chart shows a comparison of mean scores on a scale of 10 to 90. For example, respondents from 11 health centers had a higher perception score on patient-provider communication than other dimensions with less variation, while the information provision dimension was mostly ranked lowest with more variability.

**Figure 1:**
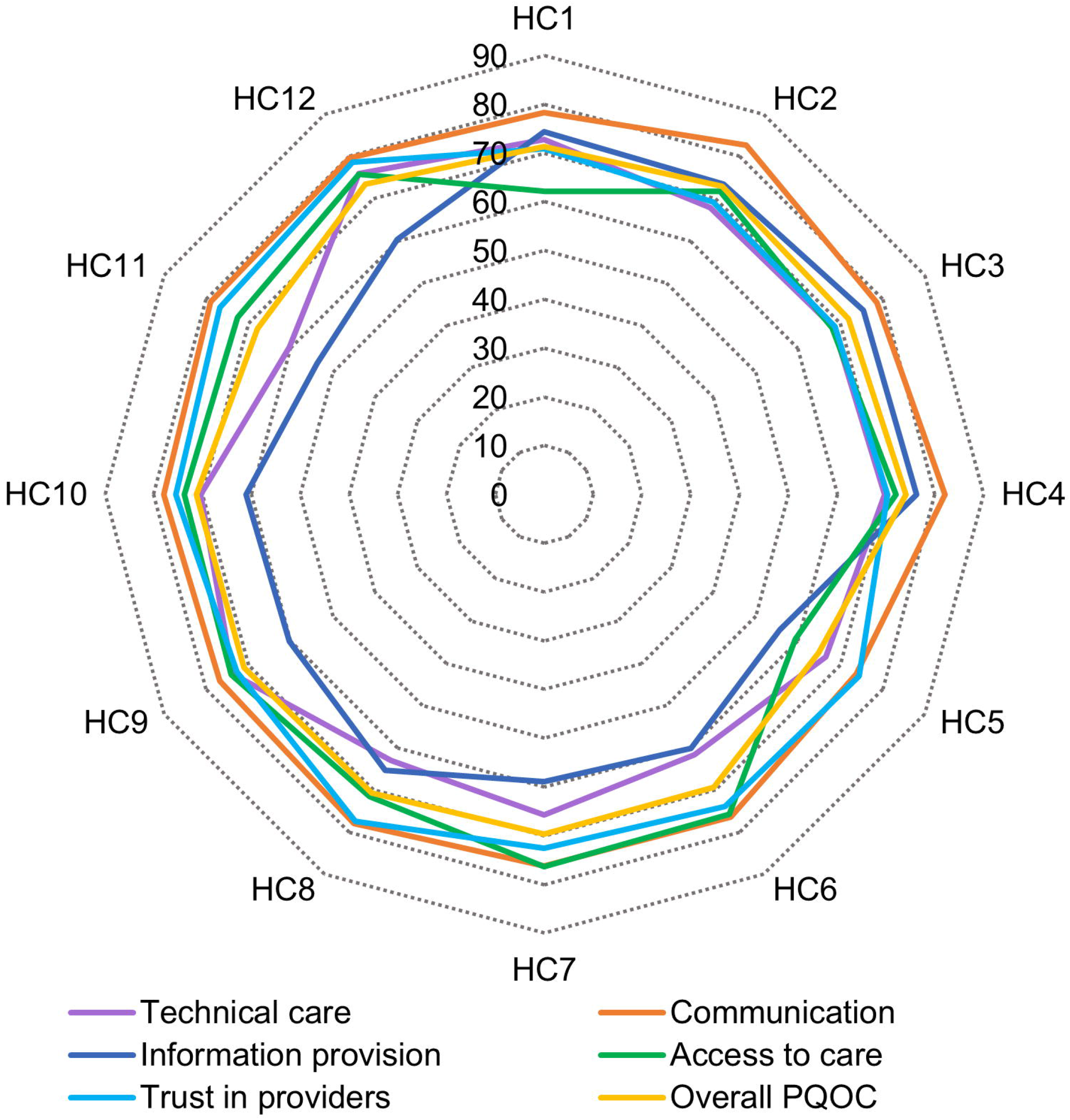
Summary of the mean scores of the PQoC and its dimensions across 12 health centers in two districts of northeast Ethiopia, 2021

### Predictors of perceived quality of care: Multilevel analysis

The fixed effects (measures of association) and the random effects (measures of variation) for the multilevel linear regression model are depicted in Table 3. In the null model, 8.5% of the total variance in PQoC was attributed to cluster-level variables. The variability between clusters was statistically significant (*τ*=5.90, p<0.001). Furthermore, the null model shows a significant improvement in fit relative to a standard linear model, demonstrating the importance of developing a multilevel model. The cluster-level variation in Model II remained significant (τ=6.33, p<0.001), with 9.31% of the total variability attributed to differences across clusters. The PCV was negative in this model, indicating that individual-level characteristics did not play a role in explaining the between cluster variation. In Model III, cluster-level variables accounted for just 1.33% of the variation in PQoC across clusters. The PCV showed that cluster-level variables explained 85.42% of the between health centers variation, indicating the importance of including cluster-level characteristics to build a more robust explanatory model. We interpreted the results of the regression analysis using Model IV, which has the lowest DIC and AIC.

**Table 3:**
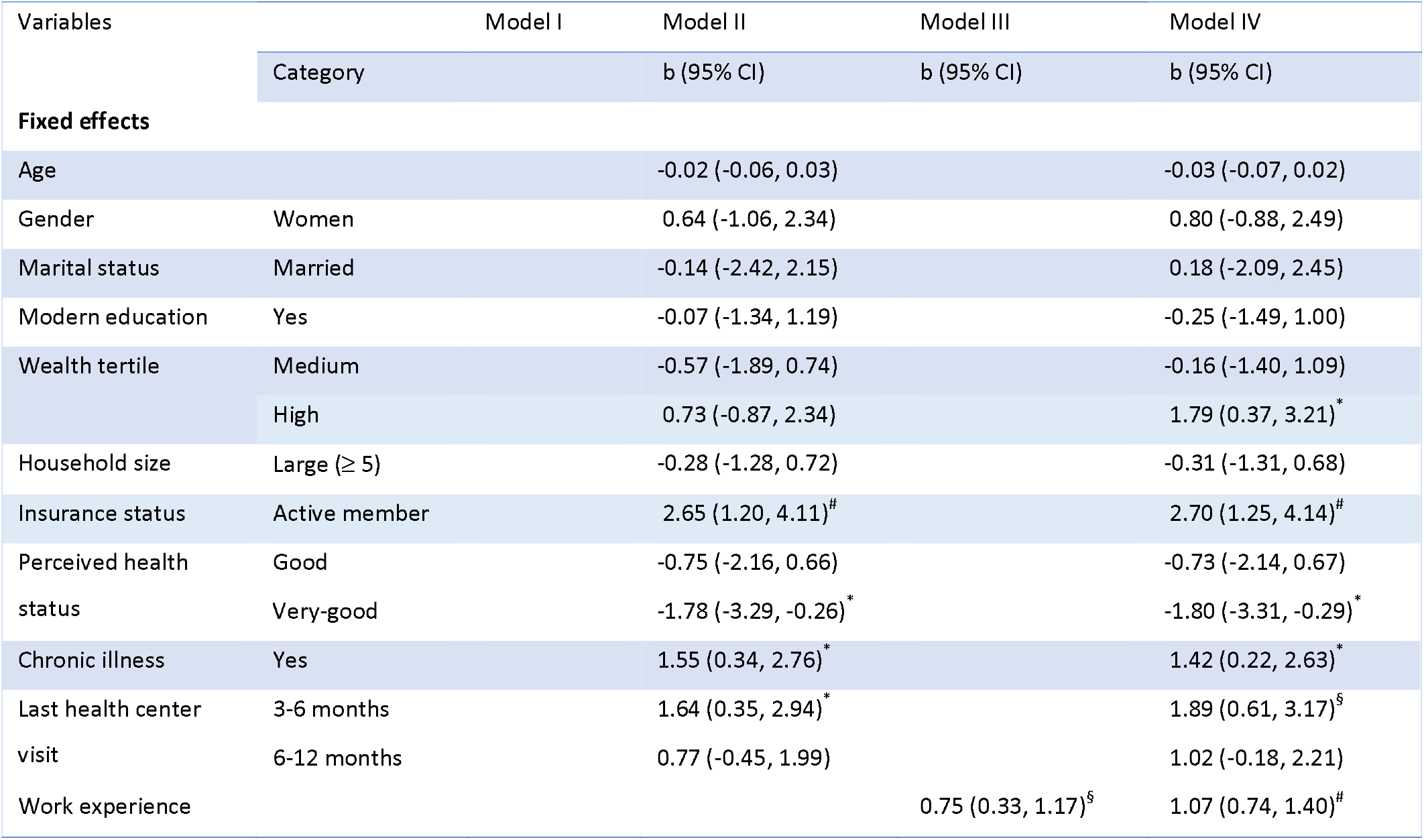
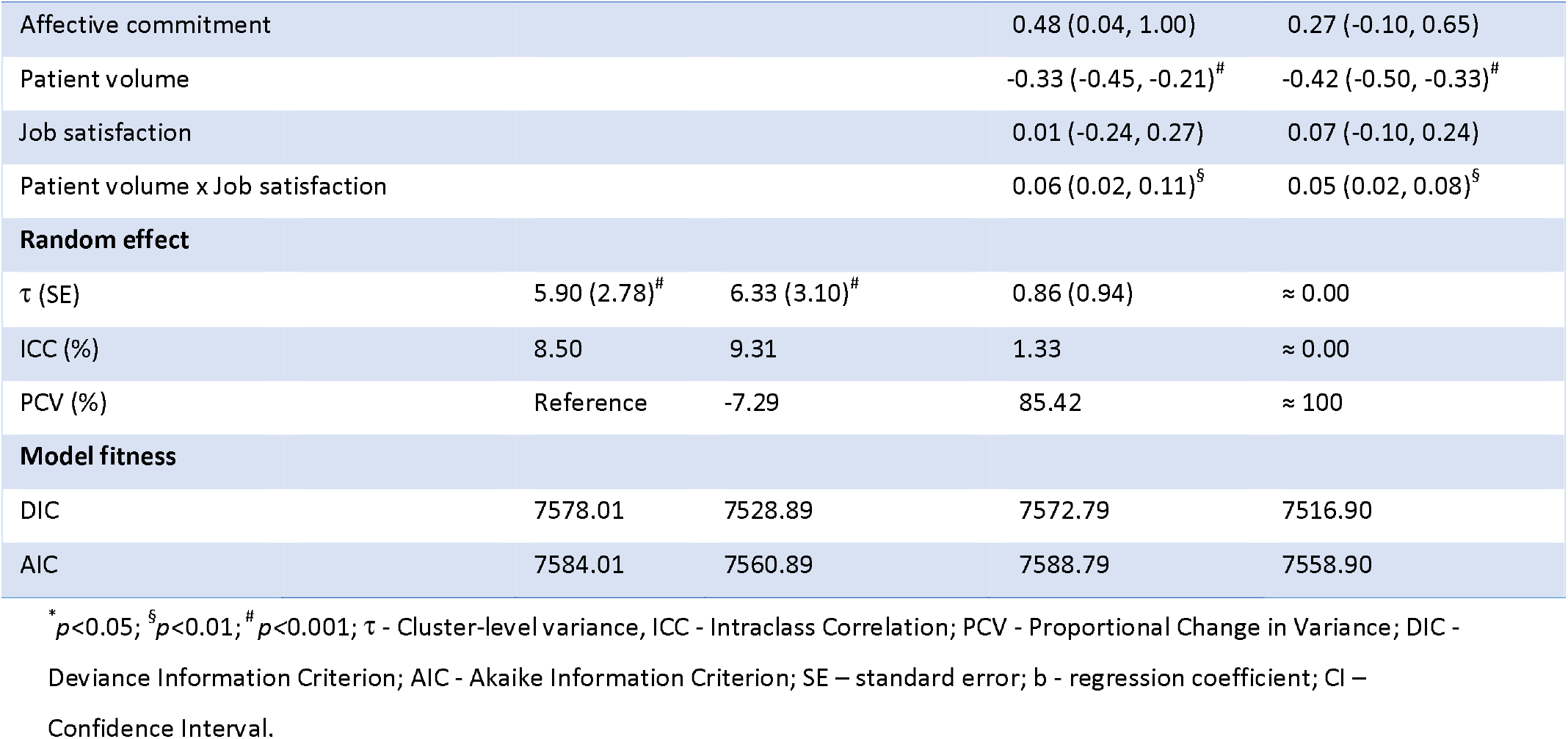
Multilevel linear regression analysis of factors associated with PQoC among households ever enrolled in a CBHI scheme in two districts of northeast Ethiopia, 2021

After adjusting for other individual and cluster-level factors, the mean PQoC score for households with higher wealth tertile increased by 1.79 points compared to those with lower wealth tertile (b=1.79; 95% CI: 0.37, 3.21). Households who were active members of CBHI at the time of the study had a 2.70-point higher PQoC score than ex-members (b=2.70; 95% CI: 1.25, 4.14). The PQoC score of households who rated their health status as very-good was 1.80 points lower compared to those who rated it as fair (b=-1.80; 95% CI: -3.31, -0.29). Compared to households without a chronic illness, those with one or more family members with a chronic illness had a 1.42 point higher perception score (b=1.42; 95% CI: 0.22, 2.63). Time to a recent visit to a health center was also significantly associated with PQoC score. The mean score for households who had their most recent visit to a health center before 3-6 months was 1.89 points higher compared to those whose recent visit was within 3-months prior to the study (b=1.89; 95% CI: 0.61, 3.17).

Regarding cluster-level variables, the average work experience of health care providers and patient volume had statistically significant associations with PQoC. A 1.07-point improvement in the average PQoC score of health centers was noted for every year increase in the median work experience of health care providers (b=1.07; 95% CI: 0.74, 1.40). An interaction term between patient volume and job satisfaction was positively associated with PQoC, implying that increasing staff job satisfaction would buffer or lessen the effect between patient volume and PQoC. At an average staff job satisfaction, a 0.42-point drop in the average PQoC score of health centers was observed for a unit increase in patient volume (b=-0.42; 95% CI: -0.50, - 0.33). A one-unit increase in patient volume would only result in a 26% fall in average PQoC if the average job satisfaction is set one SD above the mean. This prediction was substantiated by the fact that the margins graph for patient volume showed the flattest slope for high job satisfaction. However, the buffering role is observed in health centers with an average patient volume of 30.75 or higher.

## DISCUSSIONS

If insured households consider the quality of care they receive under health insurance is optimal, they will maintain their membership. ^18 19^ In this study, the mean PQoC score was 70.28 from a scale of 20-100 with an SD of 8.39. The patient-provider communication received the highest score (M=77.84, SD=10.12) among the five quality dimensions. In 2015, the Ethiopian government incorporated the development of caring, respectful and compassionate health care providers as one of the main transformation agendas in its five-year strategic plan.^27^ Our finding may be attributed partly to the government’s ongoing training initiative aimed at producing caring, respectful and compassionate health care providers. The perception score for the information provision dimension, on the other hand, was the lowest (M=64.67, SD=13.87).

This could be attributed to an increase in patient volume following the implementation of CBHI.^26^ Items loaded under this dimension appear less practical in the presence of a larger patient load. If health care providers are required to treat a large number of patients, consultation times will be reduced. They are unlikely to provide the necessary information to their clients if they are under time constraints. Regarding item level observations, waiting time and medicine availability received the lowest perception scores (62.96 and 63.50, respectively), which could also be related to increased patient load. This is consistent with previous studies in Ethiopia, which showed insured clients frequently complain about a lack of medicine and long wait times at CBHI-affiliated health facilities.^42 43^

Results of the regression analysis revealed that households with higher wealth tertile had a higher PQoC score than those with lower wealth tertile. This is in contrast to other studies,^15 44^ whereby the richest group had a lower perception score. This discrepancy could be attributed to the use of different metrics to assess quality of care. People with higher economic status may be more aware of health issues and able to bargain with health care providers to obtain the best possible care. Furthermore, if prescribed medicines are not available in CBHI-affiliated health facilities (which is one of the lowest-rated items in this study), they can afford to buy from private pharmacies. On the contrary, it may be irritating for people with lower economic status to buy medicines with limited money or to forgo treatment due to lack of money. In this regard, they may develop a negative perception of the quality of care.

Households who were active members of CBHI at the time of the study had a higher rating of PQoC compared to ex-members. Contrary to our finding, a study in Ghana showed that previously insured clients had a higher perception of quality of care compared to actively insured clients (statistical significance is not reported). The authors argue this was due to the more time-consuming nature of the service delivery processes for insured clients.^45^ At least three possible explanations exist for the relationship between CBHI status and PQoC. First, because they do not have to pay for health care, active members have better access to and enjoy its benefits, resulting in a favorable perception of its quality. Second, the relationship could be due to an endogeneity issue. It is plausible that higher quality score reported by active members is due to their desire to stay in the scheme, which could be influenced by unobserved variables. We tested for endogeneity between current insurance status and PQoC using the Durbin–Wu–Hausman test, and the results showed no evidence of endogeneity. However, there is still the possibility of endogeneity due to omitted variables. Third, ex-members of CBHI may have had negative experiences with health services, which led to the decision to discontinue their membership. As a result, they would be critical in rating the quality of care provided. In support of the latter argument, it was evidenced that poor quality of care was a major reason for insurance members to leave the scheme.^24 46^ Elsewhere, a statistically significant association was also reported between dropout and low quality of care.^47 48^

This study verified that the PQoC score of households who rated their health status as very-good was significantly lower compared to those who rated it as fair. The households’ chronic illness experiences also influence the PQoC rating. The PQoC score of households with a chronic illness was higher compared to those without a chronic illness. This may be true for people who perceive their health as fair or who live with chronic conditions to appreciate the gains or benefits of the health care they received. In this respect, they may be more likely than their counterparts to rate quality of care higher.

The results also indicated that households who had their most recent visit to a health center before 3-6 months had higher PQoC scores compared to those whose recent visit was within 3-months prior to the study. Patients may experience varying levels of emotional highs and lows, depending on the length of the most recent facility visit. Although patients’ perceptions of quality may develop over time,^5^ patients who recently visited a health facility may be more critical of the quality of care due to strong emotions attached to negative events or health services that fall short of their expectations.

Our findings revealed that the average work experience of health care providers was positively associated with PQoC. Work experience is linked to task specialization, which can lead to a faster work pace, more output in less time, and higher quality.^49^ This could be more pronounced in Ethiopia, where the number of outpatient visits to CBHI-affiliated health centers had increased dramatically.^26^ Providers with more experience take less time to make diagnoses and treatment decisions, while still providing recommended practical aspects of care, such as good communication, physical examination, and provision of relevant health information.^49^ As a result, they can reduce waiting times, and their management outcomes may be more effective than inexperienced providers.

Conditional on the average staff job satisfaction, patient volume has a negative association with PQoC. A study in Ethiopia identified a non-linear significant association (an inverted U-shape) between patient volume and quality. Quality decreased with increasing patient volume in health facilities that treated 90.6 or more patients per day, while quality increased with increasing patient volume in health facilities that treated less than 90.6 patients per day in the outpatient departments.^50^ Our finding is consistent with a study at public hospitals in China,^30^ where overcrowding was negatively associated with clients’ perception of quality of care. There are two possible explanations for the observed relationship between patient volume and PQoC. First, increased patient volume would put a great deal of pressure on health care providers to treat a large number of patients in a short time. This may result in shorter consultation time and the omission of important practical aspects of care. Second, an increase in patient volume would mean longer waiting times at various service delivery points. Both these factors could have contributed to a negative patient experience and influenced their perception on overall quality of care. Some studies reported a positive relationship between patient volume and quality of basic maternal care, and postoperative infections.^51 52^ The alternative direction of this relationship, in which quality drives patient volume, is based on the assumption that the provision of high-quality care will attract more patients. This may be true in areas where patients have access to competitive health care facilities, and health care providers are incentivized for providing higher quality care. This is not the case in low-income countries, like Ethiopia, where health care facilities are hard to reach for most rural populations. Members of CBHI are further limited to use health services only in public health facilities affiliated with the scheme.

This study found no relationship between staff job satisfaction and PQoC. This contrasts with the findings of Kvist et al,^53^ which reported a positive relationship between job satisfaction among nursing staff and patients’ perceptions of quality of care. Despite this, it moderates the relationship between patient volume and PQoC in a nonlinear fashion. Increased job satisfaction buffers the negative relationship between patient volume and PQoC in health centers with an average patient volume of 30.75 or higher. When the average patient volume is less than 30.75, however, an increase in job satisfaction enhances the effect between patient volume and PQoC. It is plausible that the buffering role of provider job satisfaction as patient volume rises indicates that provider job satisfaction is a result of the intrinsic rewards of higher work performance. Providers may also be fully available during working hours at the health facility due to the increased number of clients. On the other hand, the moderating role of enhancing the relationship as patient volume decreases could suggest that a low workload is one source of job satisfaction. Because clients are in small numbers, providers may not be fully engaged during working hours. They may have the freedom to do other businesses outside the health facility, leaving patients unattended and dissatisfied.

The findings of this study will be an essential input for quality improvement initiatives as well as addressing challenges in the country’s efforts to establish higher-level insurance pools. This is the first study of its kind to consider cluster-level variables associated with PQoC in Ethiopia. It gives an important lesson to health care managers and other relevant stakeholders to consider cluster-level characteristics in healthcare quality improvement efforts. It also pointed out quality dimensions that require special consideration in managerial decisions. Despite the significant findings of the current study, some caution should be taken in interpreting the findings. One noteworthy limitation of this study is the cross-sectional nature of the data. The study’s analysis was conducted to identify for associations rather than prove causation. Second, the association between current insurance status and PQoC could be due to the possibility of endogeneity. Third, patient volume data based on secondary data may not reflect the true figure due to the possibility of under or over reporting.

## CONCLUSIONS

Despite encouraging findings on patient-provider communication, much work remains to be done to improve information provision and access to care quality dimensions. According to the findings, people’s perceptions of quality of care varied depending on a variety of individual and cluster-level factors. The household’s wealth status, current insurance membership, perceived health status, presence of chronic illness in the household, and time to a recent visit to a health center were individual-level predictors of PQoC. At the cluster-level, patient volume and work experience of health care providers were associated with PQoC. A lower patient volume allows the health care provider to devote more time and attention to each patient, address their patients’ individual needs, and have more time to improve communication with and provide behavior change counseling, which has an impact on quality of care.^54^ Therefore, to ensure that patients have access to a better quality of care, it is critical to determine an appropriate patient volume per care provider. Staff job satisfaction was an important factor that buffers the effect between patient volume and PQoC. Hence, it is vital to devise mechanisms to improve staff job satisfaction, especially in health facilities with higher patient volume. More importantly, health centers should go to great lengths to ensure that every patient has access to the necessary medications. This will boost clients’ trust in health care providers, which will be critical for health insurance schemes to retain and attract members.

## Data Availability

Data are available in a public, open access repository. The datasets generated and analyzed during the current study are available in the Dryad repository.

https://doi.org/10.5061/dryad.ncjsxksw5

## Acknowledgements

The authors would like to acknowledge the health offices of Tehulederie and Kallu districts, health extension workers, Kebele leaders, data collectors, supervisors, and study participants. I (MH) want to acknowledge Bahir Dar university for the opportunity it has given me to pursue my PhD study.

## Contributors

MH conceptualized the study, designed the study, collected the data, analyzed and interpreted the data, and drafted the manuscript. MA and NBB contributed to survey design, data collection and statistical analysis, and reviewed the manuscript. All authors read and approved the final manuscript.

## Patient consent for publication

Not required.

## Ethics approval

Ethical approval was obtained from the Institutional Review Board (IRB) of College of Medicine and Health Science, Bahir Dar University with protocol number 001/2021. A support letter was communicated to the district health offices to gain entry permission into the community where the research was conducted. Before the interview, verbal informed consent was secured from each of the study participants. Confidentiality was assured through collecting anonymous information and informing the participants that personal identifiers would not be revealed to a third party.

## Provenance and peer review

Not commissioned; externally peer reviewed.

## Data availability statement

Data are available in a public, open access repository. The datasets generated, and analyzed during the current study are available in the Dryad repository, at https://doi.org/10.5061/dryad.ncjsxksw5

## Open access

This is an open access article distributed in accordance with the Creative Commons Attribution Non-Commercial (CC BY-NC 4.0) license, which permits others to distribute, remix, adapt, build upon this work non-commercially, and license their derivative works on different terms, provided the original work is properly cited, appropriate credit is given, any changes made indicated, and the use is non-commercial. See: http://creativecommons.org/licenses/by-nc/4.0/.

