## Supplementary file 1 for "Perceived quality of care among households ever enrolled in a community-based health insurance scheme in two districts of northeast Ethiopia: a multilevel analysis"

Supplementary file 1: Factor analysis of the measurement scale to assess the perceived quality of care among households enrolled in a CBHI in two districts of northeast Ethiopia, 2021

| Dimensions and items | Loadings under each dimension | | | | |
|---|---|---|---|---|---|
| | 1 | 2 | 3 | 4 | 5 |
| **Technical care** | | | | | |
| The necessary Laboratory tests were performed | 0.911 | | | | |
| Health care providers perform the necessary physical examinations | 0.818 | | | | |
| Health care providers make a good diagnosis | 0.740 | | | | |
| **Patient-provider communication** | | | | | |
| Health care providers actively ask questions to understand your situation | | 0.846 | | | |
| Health care providers listened to you carefully what you had to say | | 0.845 | | | |
| Health care providers treated you with courtesy and respect | | 0.542 | | | |
| **Information provision** | | | | | |
| Health care providers clearly explained the use and side effects of medicines | | | 0.787 | | |
| Health care providers clearly explained the results of tests and examination | | | 0.760 | | |
| Health care providers explain things in a way you could understand | | | 0.672 | | |
| Health care providers spent sufficient time examining patients | | | 0.510 | | |
| **Access to care** | | | | | |
| Patients do not wait long in the health center to receive treatment | | | | 0.799 | |
| All prescribed medicines are available on the spot | | | | 0.624 | |
| Facility assistants are friendly and helpful to patients | | | | 0.559 | |
| The health facility serves all patients fairly | | | | 0.463 | |
| **Trust in care providers** | | | | | |
| Treatment is effective for recovery and cure | | | | | 0.754 |
| Health care providers prescribe appropriate medicines for patients | | | | | 0.672 |
| You have confidence in the competence of health care providers | | | | | 0.662 |
