## Supplementary file 2 for "Perceived quality of care among households ever enrolled in a community-based health insurance scheme in two districts of northeast Ethiopia: a multilevel analysis"

Supplementary file 2: Mean score of each measurement item of the perceived quality of care (20-100 scale) among households enrolled in a CBHI in two districts of northeast Ethiopia, 2021

| **Factors and items** | | | **95% CI** | |
|---|---|---|---|---|
| | **Mean** | **SD** | **LCI** | **UCI** |
| **Technical care** | **68.34** | **15.24** | **67.43** | **69.25** |
| The necessary Laboratory tests were performed | 69.20 | 18.36 | 68.10 | 70.29 |
| Health care providers perform the necessary physical examinations | 68.23 | 18.89 | 67.11 | 69.36 |
| Health care providers make good diagnosis | 67.59 | 17.69 | 66.53 | 68.64 |
| **Patient-provider communication** | **77.84** | **10.12** | **77.23** | **78.44** |
| Health care providers actively ask questions to understand your situation | 80.39 | 11.68 | 79.69 | 81.09 |
| Health care providers listened to you carefully what you had to say | 79.61 | 10.93 | 78.96 | 80.26 |
| Health care providers treated you with courtesy and respect | 73.51 | 16.72 | 72.51 | 74.50 |
| **Information provision** | **64.67** | **13.87** | **63.84** | **65.49** |
| Health care providers clearly explained the use and side effects of medicines | 62.90 | 19.87 | 61.72 | 64.09 |
| Health care providers clearly explained the results of tests and examination | 62.50 | 19.48 | 61.34 | 63.66 |
| Health care providers explain things in a way you could understand | 69.36 | 17.42 | 68.32 | 70.40 |
| Health care providers spent sufficient time to examine patients | 63.90 | 20.18 | 62.70 | 65.11 |
| **Access to care** | **69.47** | **11.77** | **68.77** | **70.17** |
| Patients do not wait long in the health center to receive treatment | 62.96 | 20.17 | 61.76 | 64.16 |
| All prescribed medicines are available on the spot | 63.50 | 20.37 | 62.28 | 64.71 |
| Facility assistants are friendly and helpful to patients | 73.38 | 16.07 | 72.42 | 74.34 |
| The health facility serves all patients fairly | 78.06 | 15.90 | 77.11 | 79.01 |
| **Trust in care providers** | **73.20** | **11.02** | **72.55** | **73.86** |
| Treatment is effective for recovery and cure | 72.47 | 14.78 | 71.59 | 73.35 |
| Health care providers prescribe appropriate medicines for patients | 75.47 | 12.90 | 74.70 | 76.24 |
| You have confidence in the competence of health care providers | 71.67 | 14.36 | 70.82 | 72.53 |
| **Overall perceived quality of care (PQoC)** | **70.28** | **8.39** | **69.77** | **70.78** |
